# Replication of LZTFL1 gene region as a susceptibility locus for COVID-19 in Latvian population

**DOI:** 10.1101/2021.03.31.21254708

**Authors:** Raimonds Rescenko, Raitis Peculis, Monta Ustinova, Laura Ansone, Anna Terentjeva, Helena Daiga Litvina, Liga Birzniece, Kaspars Megnis, Oksana Kolesova, Baiba Rozentale, Ludmila Viksna, Vita Rovite, Janis Klovins

## Abstract

The severity of COVID-19 disease is partly determined by host genetic factors that have been reported by GWAS. We evaluated nine previously reported genome-wide significant associations regardless of the disease severity in a representative sample from the population of Latvia. Our cohort consisted of 475 SARS-CoV-2 positive cases, from which 146 were hospitalized individuals and 2217 controls. We found three variants from Neanderthal introgression event at the 3p21.31 region to be significantly associated with increased risk of SARS-CoV-2 infection and hospitalization status. The strongest association was displayed by rs71325088 with Bonferroni adjusted P=0.007, OR=1.46 [95% CI 1.17-1.81]. We performed fine-mapping by exploring 1 Mb region at 3p21.31 locus and identified 9 SNPs with even lower p-values with the strongest association estimated for rs2191031 P=5e-05, OR = 1.40[CI 95% 1.19-1.64] located in the LZTFL1. We show clear replication of 3p.21.31 locus in an independent cohort which favors further functional investigation of leading variants.

## Main text

Clinical outcomes of Sars-Cov-2 infection are highly heterogeneous, ranging from symptoms-free infection to death of a patient (Park et al., 2020). Although the outcome can be explained to some extent by age and comorbidities, genetic factors have also been linked to the prognosis of SARS-CoV-2 infection (Hu et al., 2021; Liu et al., 2021; Maya et al., 2020; Pairo-Castineira et al., 2021; The Severe Covid-19 GWAS Group, 2020; Zeberg and Pääbo, 2020). To this date, the association of nine single nucleotide polymorphisms (SNPs) at genome-wide significance level has been reported in the GWAS catalog for severe COVID-19 outcome phenotype (MONDO_0100096) (GWAS catalog, 2021). These SNPs were identified in two studies involving subjects from populations of diverse Eurasian and American origin (Pairo-Castineira et al., 2021; The Severe Covid-19 GWAS Group, 2020). A leading association across these studies is found in the 3p21.31 region, specifically near genes *LZTFL1* and *SLC6A20* (Pairo-Castineira et al., 2021; The Severe Covid-19 GWAS Group, 2020) and rs11385942 variant from this region also shows the lowest heterogenous P-value in a worldwide cohort of 49 studies from 19 countries (The COVID-19 Host Genetics Initiative and Ganna, 2021).

While the primary attention in previous studies was to identify genetic associations with the severity of the COVID-19 disease, we decided to evaluate already reported genome-wide significant associations regardless of the disease severity in a representative sample of 2692 individuals from the population of Latvia. Additionally, we aimed to further examine the corresponding loci to identify new associated polymorphisms in a region with positive signals. The study included all 475 COVID-19 patients recruited to the Genome Database of Latvian Population (LGDB) (Rovite et al., 2018) from June 2020 to January 2021. This cohort consisted of 146 hospitalized patients, while the rest represented patients with less severe symptoms or asymptomatic cases. In total, 2217 controls were selected from LGDB, representing the group of the general population and disease-specific biobank participants from the Latvian population with available genome-wide genotyping data. This study was approved by the Central Medical Ethics Committee of Latvia (No. 01-29.1.2/928). The mean age in cases and controls were 47.3 (SD = 17.8) and 54.9 (SD = 13.1) respectively. The proportion of females was 64.5% in the case group and 62.0 % in the controls. After the genotype quality control and imputation using the TOPMed r2 imputation server, we performed an association study with sex and age as covariates. Association was performed with PLINK 1.9 (Chang et al., 2015) according to the parameters defined in SAIGE software (Zhou et al., 2020), adding the first 20 PCs to control for population stratification.

We selected a total of nine significantly associated SNPs reported in the GWAS catalog from two previously published studies analyzing the association between severe COVID-19 cases and population controls (Pairo-Castineira et al., 2021; The Severe Covid-19 GWAS Group, 2020). Table 1 summarizes the list of the selected SNPs. No significant deviation in allele frequencies (AF) was found between the control group and the global AFs from the 1000G Phase 3 reference set (The 1000 Genomes Project Consortium, Auton, et al., 2015). Out of the nine selected variants, we identified three significantly associated *LZTFL1* gene polymorphisms rs71325088 (0.007, OR 1.46 [1.17-1.81]), rs11385942 (0.005, OR 1.47 [1.18-1.82]) and rs73064425 (0.007, OR 1.45 [1.17-1.80]) after applying the Bonferroni correction for multiple testing (Table 1). We observed a high degree of linkage disequilibrium (LD) between all three SNPs reflected by almost identical frequencies in case and control groups. These variants were also the most significant (Phet = 7.2e-25) in the worldwide meta-analysis of individuals with the SARS-CoV-2 infection, hospitalization, and critical illness (The COVID-19 Host Genetics Initiative and Ganna, 2021). All of these variants are believed to represent the region to be a remnant of Neanderthal gene pool introgression into the modern human population (Zeberg and Pääbo, 2020). *LZFTL1* gene has been implicated in ciliogenesis and intracellular trafficking of ciliary proteins, probably impacting airway epithelial cell function (Promchan et al., 2020; Shelton et al., 2021). Even though the primary analysis was performed on the case group that included all patients regardless of severity, there is an obvious bias toward the inclusion of symptomatic patients as they are more likely to be tested for the SARS-CoV-2 presence than asymptomatic cases. We also tested the association of the selected SNPs with disease severity in our study group using hospitalized patients as cases against the same group of controls (Table 1).

**Table 1.** Association of selected single nucleotide polymorphisms and SARS-CoV-2 infection

| Chr | SNP ID | Risk allele | Mapped genes | Covid-19 patients (n=475)<br>vs. controls (n=2217) |  |  | Hospitalized Covid-19 patients (n=146)<br>vs. controls (n=2217) |  |  |  |
| --- | --- | --- | --- | --- | --- | --- | --- | --- | --- | --- |
|  |  |  |  | P/<br>Padj | AF<br>case/<br>cntrl | OR<br>[95% CI] | P/<br>Padj | AF<br>case/<br>cntrl | OR<br>[95% CI] | GWAS<br>catalog<br>OR |
| 3 | rs71325088 | C | SLC6A20,<br>LZTFL1 | 7e-04/<br>**0.007** | 0.13/<br>0.09 | 1.46<br>[1.17-1.81] | 1e-05/<br>**1e-04** | 0.16/<br>0.09 | 2.10<br>[1.50-2.92] | 1.9<sup>a</sup><br>[1.73-2.0] |
| 3 | rs11385942 | A | LZTFL1 | 5e-04/<br>**0.005** | 0.13/<br>0.09 | 1.47<br>[1.18-1.82] | 6e-06/<br>**6e-05** | 0.16/<br>0.09 | 2.14<br>[1.54-2.99] | 2.11<sup>b</sup><br>[1.70-2.61] |
| 3 | rs73064425 | T | AC099782.3,<br>LZTFL1 | 8e-04/<br>**0.007** | 0.13/<br>0.09 | 1.45<br>[1.17-1.80] | 1e-05/<br>**9e-05** | 0.16/<br>0.09 | 2.13<br>[1.52-2.97] | 1.7<sup>a</sup><br>NR |
| 6 | rs143334143 | A | CCHCR1 | 0.7/<br>1 | 0.09/<br>0.09 | 0.94<br>[0.73-1.22] | 0.4/<br>1 | 0.10/<br>0.09 | 1.18<br>[0.78-1.80] | 1.3<sup>a</sup><br>[1.27-1.48] |
| 9 | rs657152 | A | ABO | 0.1/<br>0.891 | 0.47/<br>0.44 | 1.13<br>[0.98-1.31] | 0.1/<br>1 | 0.48/<br>0.44 | 1.22 [0.95-1.55] | 1.32<sup>b</sup><br>[1.20-1.47] |
| 12 | rs6489867 | C | OAS1,<br>AC004551.1 | 0.9/<br>1 | 0.30/<br>0.31 | 0.99<br>[0.85-1.16] | 0.7/<br>1 | 0.29/<br>0.31 | 0.96<br>[0.73-1.25] | 1.2<sup>a</sup><br>[1.14-1.25] |
| 19 | rs2109069 | A | DPP9 | 0.01/<br>0.115 | 0.30/<br>0.27 | 1.22<br>[1.04-1.44] | 0.1/<br>1 | 0.30/<br>0.27 | 1.22<br>[0.94-1.59] | 1.2<sup>a</sup><br>[1.19-1.31] |
| 19 | rs11085727 | T | TYK2 | 0.03/<br>0.235 | 0.31/<br>0.28 | 1.20<br>[1.02-1.40] | 0.1/<br>1 | 0.32/<br>0.28 | 1.23<br>[0.95-1.60] | 1.2<sup>a</sup><br>[1.18-1.31] |
| 21 | rs13050728 | T | AP000295.1,<br>IFNAR2 | 0.6/<br>1 | 0.39/<br>0.38 | 1.04<br>[0.90-1.21] | 0.06/<br>1 | 0.43/<br>0.38 | 1.28<br>[0.99-1.64] | 1.2<sup>a</sup><br>[1.16-1.28] |

CHR, chromosome number; SNP, single nucleotide polymorphism; AF, allele frequency; Padj, Bonferroni adjusted P-value; cntrl, controls; OR, odds ratio; CI confidence interval; <sup>a</sup>, from Pairo-Castineira et al, 2020; <sup>b</sup>, The Severe Covid-19 GWAS Group, 2020

All three SNPs from the 3p21.31 region displayed a stronger association in the frame of this comparison (P=6e-05, OR=2.14 [CI 95% 1.54-2.99] and 4.2 times higher homozygote prevalence for rs11385942), supporting the notion that this locus increases the risk for respiratory symptoms. Studies that would account for participants’ exposure to the infection are needed to answer this question. Even though the other SNPs included in the study did not reach statistical significance, the odds ratios for almost all SNPs (excluding rs6489867) was similar to those previously published (Pairo-Castineira et al., 2021; The Severe Covid-19 GWAS Group, 2020) (Table 1) and probably larger sample size is needed to replicate other loci. It should be noted that the 3p.21.31 locus is the only replicated locus so far out of all 28 COVID-19 variants released in GWAS catalog to date, representing variants above P=5e-8 level.

We also explored the 500 kb regions around the rs11385942 to evaluate the association of other SNPs with the main trait in our study. In total nine 3p21.31 locus polymorphisms rs2191031, rs3774641, rs7289367, rs35896106, rs13071258, rs34668658, rs17763742, rs17763537 and rs73062389 displayed lower p-value in our cohort compared to the leading rs11385942, with the strongest association estimated for rs2191031 P=5e-05, OR = 1.40[CI 95% 1.19-1.64] located in the LZTFL1 gene (Figure 1). Rs2191031 has a 2.3 times higher prevalence in our population compared to replicated polymorphisms. To date, no function or clinical relevance has been ascribed for this variant; however, it is equivalently associated with differential expression in the esophagus mucosa (1e-5, NES -.22) (GTEx Consortium, 2018) and multiple other tissues pointing to similar etiology as replicated variants (Shelton et al., 2021). However, it is important to note that none of these nine variants had a lower P value than rs11385942 in a cohort of hospitalized COVID-19 patients.

**Figure 1.**
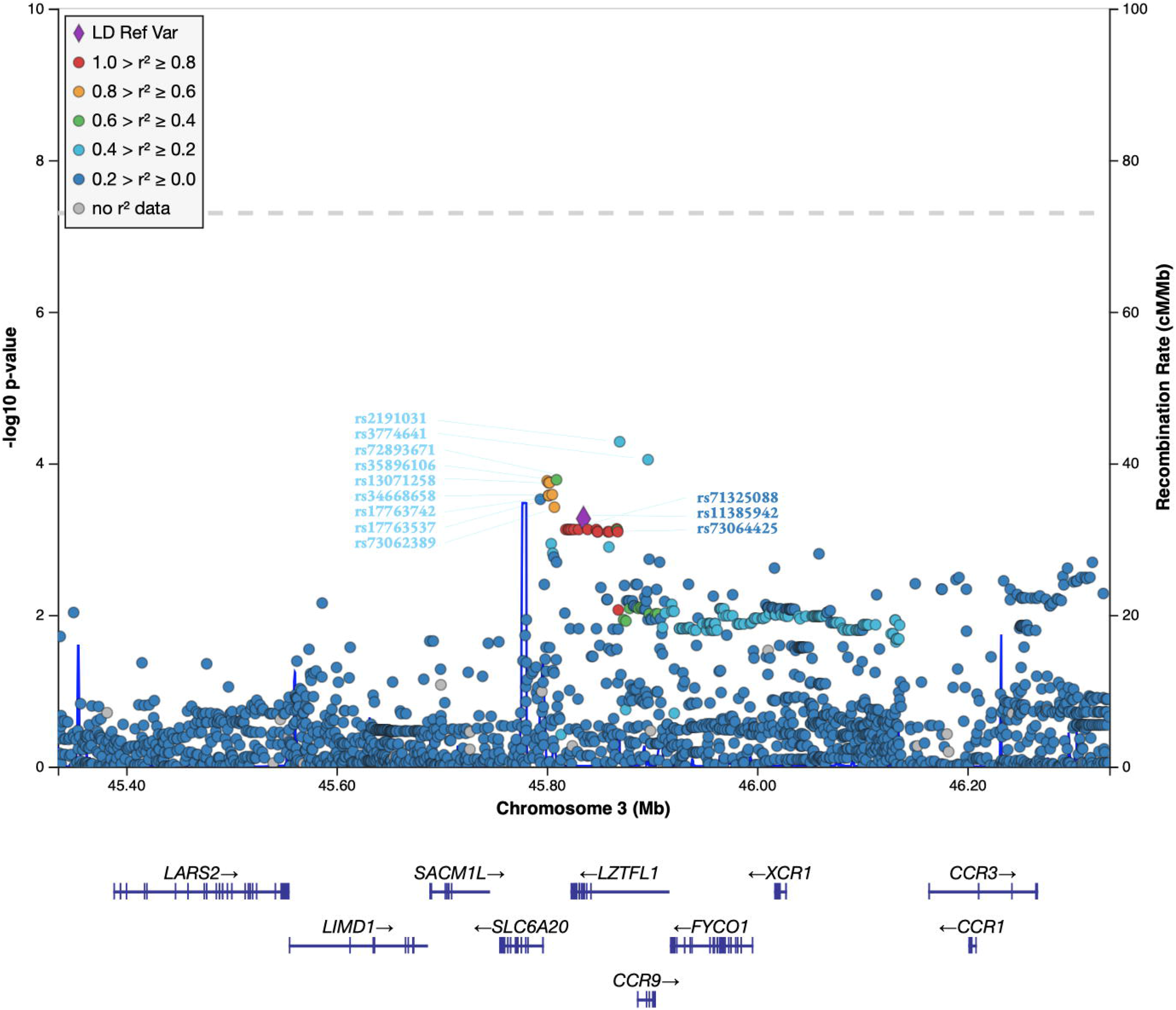
Manhattan plot of 1Mb region at 3p21.31 locus. rs11385942 variant is emphasized with a diamond shape and set as reference for LD estimation with other SNPs in the region. Rs codes for three SNPs selected for replication are depicted in dark blue, while rs codes for other SNPs displaying lower p-value in our association study are in light blue.

Using the retrospectively collected samples from a population-based biobank has some limitations. We cannot assess exposure to SARS-CoV-2 for this group of people and match that with the case group, as we do not have information on the possible rate of infection among this group. However, such a design does not increase type I error, and we did not aim to find genetic factors facilitating protection against SARS-CoV-2 infection in our study where such a design would not be appropriate. Another shortcoming is that most patients included in this study were recruited in December 2020, and extensive phenotype data, including follow-ups, are not yet available for analysis. Such data would allow elaborating interaction between different clinical data and genotype and include the development of post-COVID-19 complications as an essential phenotype for association study.

In conclusion, we demonstrate supportive evidence for the involvement of human 3p21.31 locus in the pathophysiology of COVID-19 disease using an independent cohort of patients and controls from the Latvian population. It highlights the importance of this genomic region for genetic risk estimation in relation to SARS-CoV-2 infection and the robustness of proper genetic association studies for replication purposes. Notably, the results presented here provide a preliminary indication of variants with possible functional effects and calls for further studies exploring the validation of these variants.

## Data Availability

The raw data supporting the current study are available from the corresponding author on request.

## Acknowledgments

This study was funded by the Ministry of Education and Science, Republic of Latvia, project “Establishment of COVID-19 related biobank and integrated platform for research data in Latvia”, project No. VPP-COVID-2020/1-0016. We acknowledge The Boris and Inara Teterev Foundation for support to Riga Stradins University inpatient sample collection. The authors acknowledge the Latvian Biomedical Research and Study Centre and the Genome Database of the Latvian Population for providing the infrastructure, biological material, and data.

